# Evaluation of the diagnostic value of YiDiXie™-SS, YiDiXie™-HS and YiDiXie™-D in brain malignant tumors

**DOI:** 10.1101/2024.08.18.24311913

**Authors:** Yutong Wu, Chen Sun, Zhenjian Ge, Huimei Zhou, Xutai Li, Wenkang Chen, Yingqi Li, Shengjie Lin, Pengwu Zhang, Wuping Wang, Siwei Chen, Wei Li, Jun Hu, Ling Ji, Yongqing Lai

**Author notes:** Corresponding author: Yongqing Lai, Peking University Shenzhen Hospital, 1120 Lianhua Road, Shenzhen 518036, E mail; Ling Ji, Peking University Shenzhen Hospital, 1120 Lianhua Road, Shenzhen 518036, E mail; Jun Hu, Peking University Shenzhen Hospital, 1120 Lianhua Road, Shenzhen 518036, E mail. Contributed equally to this work (Yutong Wu, Chen Sun, Zhenjian Ge, Xutai Li).

## Abstract

**Background:** Brain malignant tumors is a serious threat to human health and causes heavy economic burden. Enhanced MRI is widely used in the diagnosis of brain tumors. However, false-positive results of enhanced MRI will lead to misdiagnosis and incorrect surgery or treatment, while false-negative results of enhanced MRI will lead to underdiagnosis of malignant tumors and delayed treatment. There is an urgent need to find convenient, cost-effective and non-invasive diagnostic methods to reduce the false-positive and false-negative rates of brain-enhanced MRI. The aim of this study was to evaluate the diagnostic value of YiDiXie™-SS, YiDiXie™-HS and YiDiXie™-D in brain malignant tumors.

**Patients and methods:** 233 subjects (malignant group, n=74; benign group, n=159) were finally included in this study. Remaining serum samples from the subjects were collected and tested by applying the YiDiXie™ all-cancer detection kit to evaluate the sensitivity and specificity of YiDiXie™-SS, YiDiXie™-HS and YiDiXie™-D.

**Results:** The sensitivity of YiDiXie™ SS was 97.3% (95% CI: 90.7% - 99.5%) and its specificity was 63.5% (95% CI: 55.8% - 70.6%). This means that YiDiXie ™ -SS has a very high sensitivity and specificity in brain tumors.YiDiXie ™ -HS has a sensitivity of 83.8% (95% CI: 73.8% - 90.5%) and a specificity of 84.9% (95% CI: 78.5% - 89.6%). This means that YiDiXie™-HS has high sensitivity and specificity in brain tumors.YiDiXie™-D has a sensitivity of 70.3% (95% CI: 59.1% - 79.5%) and a specificity of 93.7% (95% CI: 88.8% - 96.5%). This means that YiDiXie™-D has high sensitivity and very high specificity in brain tumors. The sensitivity of YiDiXie™ SS in patients with positive enhancement MRI was 98.3% (95% CI: 91.1% - 99.9%) and its specificity was 62.5% (95% CI: 38.6% - 81.5%). This means that the application of YiDiXie ™ -SS reduces the false-positive rate of enhanced MRI by 62.5% (95% CI: 38.6% - 81.5%) with essentially no increase in the leakage of malignant tumors.The sensitivity of YiDiXie™-HS in patients with negative enhanced MRI was 85.7% (95% CI: 60.1% - 97.5%) and its specificity was 84.6% (95% CI: 77.8% - 89.6%). This means that the application of YiDiXie ™ -HS reduced the false-negative rate of enhanced MRI by 85.7% (95% CI: 60.1% - 97.5%). YiDiXie™-D had a sensitivity of 71.7% (95% CI: 59.2% - 81.5%) and a specificity of 93.8% (95% CI: 71.7% - 99.7%) in patients with positive enhanced MRI. This means that YiDiXie™ -D reduced the false-positive brain enhanced MRI rate by 93.8% (95% CI: 71.7% - 99.7%). YiDiXie™-D had a sensitivity of 64.3% (95% CI: 38.8% - 83.7%) and a specificity of 93.7% (95% CI: 88.5% - 96.7%) in patients with negative enhanced MRI. This means that YiDiXie ™ -D reduces the false-negative rate of enhanced MRI by 64.3% (95% CI: 38.8% - 83.7%) while maintaining high specificity.

**Conclusion:** YiDiXie ™ -SS has extremely high sensitivity and high specificity in brain tumors.YiDiXie ™ -HS has high sensitivity and high specificity in brain tumors.YiDiXie ™-D has high sensitivity and extremely high specificity in brain tumors.YiDiXie™-SS significantly reduces the false-positive rate of brain-enhanced MRIs with essentially no increase in delayed treatment of malignant tumors. The YiDiXie™-HS significantly reduces the false-negative rate of brain-enhanced MRI. the YiDiXie™-D can significantly reduce the false-positive rate of brain-enhanced MRI or significantly reduce the false-negative rate of brain-enhanced MRI while maintaining a high level of specificity. The YiDiXie™ test has significant diagnostic value in brain tumors, and is expected to solve the problems of “ high false-positive rate “ and “ high false-negative rate” of brain-enhanced MRI.

**Clinical trial number:** ChiCTR2200066840.

## INTRODUCTION

Brain malignant tumors is one of the common malignant tumors. According to the latest data, in 2022, there will be 321,476 new cases of brain malignant tumors in the world, and there will be 248,305 new deaths, accounting for 2.6% of the total number of deaths of malignant tumors in the world and ranking 12th^1^; Compared to 2020, the mortality rate of brain malignant tumors increased by 0.1% in 2022 and is still trending upward each year^2^. Gliomas are the most common primary intracranial tumors, accounting for approximately 80-85% of all malignant brain tumors^3^. Treatment of gliomas requires an integrated multidisciplinary approach that includes surgical resection, radiotherapy, systemic therapy, and supportive care, and surgery to maximize the safe removal of the tumor is the first-line treatment for gliomas^4^. High-grade gliomas are characterized by high aggressiveness, high disability, and high recurrence rates, with a median survival of less than two years^5-6^ and a 5-year survival rate of less than 10%. Early diagnosis not only improves survival outcomes and optimizes survival by treating early in the curable low-grade stage, but also reduces the heavy burden of treatment and the cost of treatment for families and society^7^. Therefore, brain malignant tumors are not only a serious threat to human health, but also a heavy burden to the patient, family, and society.

Enhanced MRI is widely used in the diagnosis of brain tumors. On the one hand, enhanced MRI can produce a large number of false-positive results. When enhanced MRI is positive, patients are usually treated with surgery or invasive therapy. False-positive enhanced MRI results mean that benign diseases are misdiagnosed as malignant tumors, and patients may have to bear the negative consequences of unnecessary mental pain, expensive surgery and examination costs, surgical trauma, and even loss of function. Therefore, there is an urgent need to find a convenient, cost-effective and noninvasive diagnostic method to reduce the false-positive rate of brain-enhanced MRI.

On the other hand, enhanced MRI can produce a large number of false-negative results. When enhanced MRI is negative, patients usually take observation and regular follow-up. Enhanced MRI false-negative results mean that malignant tumors are misdiagnosed as benign diseases, which will likely lead to delayed treatment, progression of malignant tumors, and may even develop into advanced stages. Patients will thus have to bear the adverse consequences of poor prognosis, high treatment costs, poor quality of life, and short survival. Therefore, there is an urgent need to find a convenient, economical and noninvasive diagnostic method to reduce the false-negative rate of brain-enhanced MRI.

In addition, there are some special patients who need to be extra cautious in choosing whether or not to operate, such as: smaller tumors, tumors involving important functional areas of the brain, and patients with poor general conditions. The risk of wrong surgery in these special patients is much higher than the risk of missed diagnosis. And the false-positive result of enhanced MRI means that benign diseases are misdiagnosed as malignant tumors, which will lead to misdiagnosis and wrong surgery. Therefore, there is an urgent need to find a convenient, cost-effective and noninvasive diagnostic method with high specificity to substantially reduce the false-positive rate of brain-enhanced MRI in these special patients or to significantly reduce its false-negative rate while maintaining high specificity.

Based on the detection of novel tumor markers of miRNA in serum, Shenzhen KeRuiDa Health Technology Co., Ltd. has developed an in vitro diagnostic test, YiDiXie ™ all-cancer test (hereinafter referred to as YiDiXie ™ test), which can detect multiple types of cancers with only 200 microliters of whole blood or 100 microliters of serum each time^8^. The YiDiXie ™ test consists of three different tests, YiDiXie™-HS, YiDiXie™-SS and YiDiXie™-D^8^.

The purpose of this study was to evaluate the diagnostic value of YiDiXie ™ -SS, YiDiXie ™ -HS and YiDiXie™-D in brain malignant tumors.

## PATIENTS AND METHODS

### Study design

The present work is part of the sub-study “ Evaluation of the YiDiXie ™ test for adjuvant diagnostic value in multiple tumors “ of the SZ-PILOT study (ChiMRIR2200066840).

The SZ-PILOT study (ChiMRIR2200066840) is a single-center, prospective, observational study. Participants signing a general informed consent for donation of remaining samples at the time of admission or physical examination were enrolled, and their remaining serum samples of 0.5 ml were collected for this study.

The present study was blinded. Neither the laboratory personnel performing the YiDiXie™ test nor the KeRuiDa laboratory technicians evaluating the results of the YiDiXie™ test were aware of the participants’ clinical information. The clinical experts assessing the subjects’ clinical information were also unaware of the results of the YiDiXie ™ test.

The present study was approved by the Ethics Committee of Shenzhen Hospital of Peking University and was conducted in accordance with the International Conference on Harmonization (ICH) Code of Practice for the Quality Management of Pharmaceutical Clinical Trials and the Declaration of Helsinki.

### Participants

Two groups of subjects were enrolled separately, and the subjects meeting the inclusion criteria were all consecutively included.

Initially, this study included hospitalized patients with “ suspected (solid or hematological) malignancy “ who signed a general informed consent form for donation of the remaining samples. Participants with a postoperative pathology diagnosis of “ malignant tumor “ were included in the malignant tumor group, and those with a postoperative pathology diagnosis of “benign tumor” were included in the benign tumor group. Subjects with ambiguous pathological findings were excluded from the present study. Some of the samples from the malignant tumor group were used in our previous article^8^.

Subjects failing the serum sample quality test prior to the YiDiXie™ test were excluded from the present study. For details of enrollment and exclusion, please refer to in our previous article^8^.

### Sample collection, processing

The serum samples used in the present study were taken from serum left over after a normal clinic visit, no additional blood draws were required. About 0.5 ml of serum samples were collected from the remaining serum of subjects in the Medical Laboratory and kept at -80 °C for the subsequent use in the YiDiXie™ test.

### The YiDiXie test

The YiDiXie ™ test was accomplished with YiDiXie ™ all-cancer detection kit, an in-vitro diagnostic kit developed and produced by Shenzhen KeRuiDa Health Technology Co..^8^ It determines the presence of cancer in a subject’s body by detecting the expression levels of dozens of miRNA biomarkers in the serum.^8^ Appropriate thresholds are predefined for each miRNA biomarker to ensure that high specificity is achieved for each miRNA marker, and these independent assays are integrated in a concurrent assay format to dramatically increase the sensitivity and maintain the high specificity for broad-spectrum cancers.^8^

The YiDiXie™ test consists of three differently characterized tests: YiDiXie™-HS, YiDiXie™-SS, and YiDiXie ™ -D.^8^ YiDiXie ™ -HS (YiDiXie ™ -Highly Sensitive, YiDiXie™-HS) has been developed with a balance of sensitivity and specificity.^8^ YiDiXie™-HS (YiDiXie ™ -Highly Sensitive, YiDiXie ™ -HS) was developed with a balance of sensitivity and specificity.^8^ YiDiXie™-SS (YiDiXie™-Super Sensitive, YiDiXie™-SS) dramatically increased the number of miRNA assays in order to achieve extremely high sensitivity for all clinical stages of all malignant tumor types.^8^ YiDiXie ™ -D (YiDiXie ™ -Diagnosis, YiDiXie ™ -D) significantly raises the diagnostic threshold of individual miRNA tests to achieve very high specificity for the whole spectrum of malignant tumor types.^8^

According to the instructions of the YiDiXie ™ all-cancer detection kit, the YiDiXie ™ test was performed. refer to in our previous article^8^ for the detailed procedure.

The raw test results were evaluated by the laboratory technicians of Shenzhen KeRuiDa Health Technology Co., Ltd. and the outcome of the YiDiXie ™ test was determined to be either “positive” or “negative”.^8^

### Diagnosis of enhanced MRI

The diagnostic conclusion of the enhanced MRI is considered “ positive “ or “ negative “. A positive result is determined if the diagnostic conclusion is positive, more positive, or favors a malignant tumor. Negative results are determined if the diagnosis is positive, more positive or favors a benign tumor, or if the diagnosis is ambiguous as to whether the tumor is benign or malignant.

### Extraction of clinical data

Clinical, pathological, laboratory, and imaging data in this study were extracted from the subjects’ hospitalized medical records or physical examination reports. For clinical staging, assessment was done by trained clinicians according to the AJCC staging manual (7th or 8th edition)^9-10^.

### Statistical analyses

Descriptive statistics were reported for demographic and baseline characteristics. The number and percentage of subjects in each category were calculated for categorical variables; for continuous variables, the total number of subjects (n), mean, standard deviation (SD) or standard error (SE), median, first quartile (Q1), third quartile (Q3), and minimum and maximum values were calculated. Wilson (score) method was used to calculate 95% confidence intervals (CI) for multiple indicators.

## RESULTS

### Participant disposition

233 subjects (malignant group, n=74; benign group, n=159) were finally included in this study. The demographic and clinical characteristics of the 233 study subjects are presented in Table 1.

**Table 1.** Participants’ demographic and clinical manifestation.

|  | Malignant (n = 74) | Benign (n = 159) | Total (N = 233) |
| --- | --- | --- | --- |
| Age, years |  |  |  |
| Mean (SD) | 44.0 (15.33) | 47.0 (12.82) | 46.0 (13.71) |
| Median (Q1,Q3) | 41 (32, 56) | 47 (37, 56) | 46 (36, 56) |
| Min, max | 6, 79 | 11, 75 | 6, 79 |
| Age, group, n (%) |  |  |  |
| < 50 | 49 (66.2) | 91 (57.2) | 140 (60.1) |
| ≥ 50 | 25 (33.8) | 68 (42.8) | 93 (39.9) |
| < 65 | 65 (87.8) | 144 (90.6) | 209 (89.7) |
| ≥ 65 | 9 (12.2) | 15 (9.4) | 24 (10.3) |
| Sex, n (%) |  |  |  |
| Female | 38 (51.4) | 58 (36.5) | 96 (41.2) |
| Male | 36 (48.6) | 101 (63.5) | 137 (58.8) |
| Body mass index (kg/m<sup>2</sup>) |  |  |  |
| n | 54 | 119 | 173 |
| Mean (SD) | 23.4 (3.57) | 23.8 (3.45) | 23.7 (3.48) |
| Median (Q1,Q3) | 23.4 (21.3, 25.7) | 23.5 (21.9, 25.8) | 23.5 (21.6, 25.8) |
| Min, max | 15.7 31.3 | 15.6 38.3 | 15.6 38.3 |
| Body mass index category, n (%) |  |  |  |
| Underweight | 5 (6.8) | 4 (2.5) | 9 (3.9) |
| Benign | 28 (37.8) | 64 (40.3) | 92 (39.5) |
| Overweight | 14 (18.9) | 42 (26.4) | 56 (24.0) |
| Obese | 7 (9.5) | 9 (5.7) | 16 (6.9) |
| Missing | 20 (27.0) | 40 (25.2) | 60 (25.8) |

Q1,Q3, first quartile, third quartile; SD, standard deviation.

The two groups of study subjects were comparable in terms of demographic and clinical characteristics (Table 1). The mean (standard deviation) age was 46.0(13.71) years and 41.2%(96/233) were female.

### Diagnostic performance of YiDiXie™-SS

As shown in Table 2, the sensitivity of YiDiXie™ SS was 97.3% (95% CI: 90.7% - 99.5%) and its specificity was 63.5% (95% CI: 55.8% - 70.6%). This means that YiDiXie™-SS has a very high sensitivity and specificity in brain tumors.

**Table 2.** The performance of YiDiXie™ -SS.

| Table 2. The performance of YiDiXie™-SS |  |  |  |
| --- | --- | --- | --- |
|  | Malignant | Benign | Total |
|  | 74 | 159 | 233 |
| Positive | 72 | 58 | 130 |
| Negative | 2 | 101 | 103 |
| SEN = 72/74 = 97.3% (90.7% - 99.5%) SPE = 101/159 = 63.5% (55.8% - 70.6%) |  |  |  |
| SEN, Sensitivity. SPE, Specificity. Two-sided 95% Wilson confidence intervals were calculated. |  |  |  |

### Diagnostic performance of YiDiXie™-HS

As shown in Table 3, YiDiXie ™ -HS has a sensitivity of 83.8% (95% CI: 73.8% - 90.5%) and a specificity of 84.9% (95% CI: 78.5% - 89.6%). This means that YiDiXie ™ -HS has high sensitivity and specificity in brain tumors.

**Table 3.** The performance of YiDiXie™-HS.

| Table 3. The performance of YiDiXie™-HS |  |  |  |
| --- | --- | --- | --- |
|  | Malignant | Benign | Total |
|  | 74 | 159 | 233 |
| Positive | 62 | 24 | 86 |
| Negative | 12 | 135 | 147 |
| SEN = 62/74 = 83.8% (73.8% - 90.5%) SPE = 135/159 = 84.9% (78.5% - 89.6%) |  |  |  |
| SEN, Sensitivity. SPE, Specificity. Two-sided 95% Wilson confidence intervals were calculated. |  |  |  |

### Diagnostic performance of YiDiXie™-D

As shown in Table 4, YiDiXie ™ -D has a sensitivity of 70.3% (95% CI: 59.1% - 79.5%) and a specificity of 93.7% (95% CI: 88.8% - 96.5%). This means that YiDiXie ™ -D has high sensitivity and very high specificity in brain tumors.

**Table 4.** The performance of YiDiXie™-D.

|  | Malignant | Benign | Total |
| --- | --- | --- | --- |
|  | 74 | 159 | 233 |
| Positive | 52 | 10 | 62 |
| Negative | 22 | 149 | 171 |

SEN = $52/74 = 70.3\%$ (59.1% - 79.5%)
SPE = $149/159 = 93.7\%$ (88.8% - 96.5%)
SEN, Sensitivity. SPE, Specificity. Two-sided 95% Wilson confidence intervals were calculated.

### Diagnostic performance of enhanced MRI

As shown in Table 5, the sensitivity of enhanced MRI was 81.3% (95% CI: 71.1% - 88.5%) and its specificity was 90.0% (95% CI: 84.4% - 93.8%).

**Table 5.**
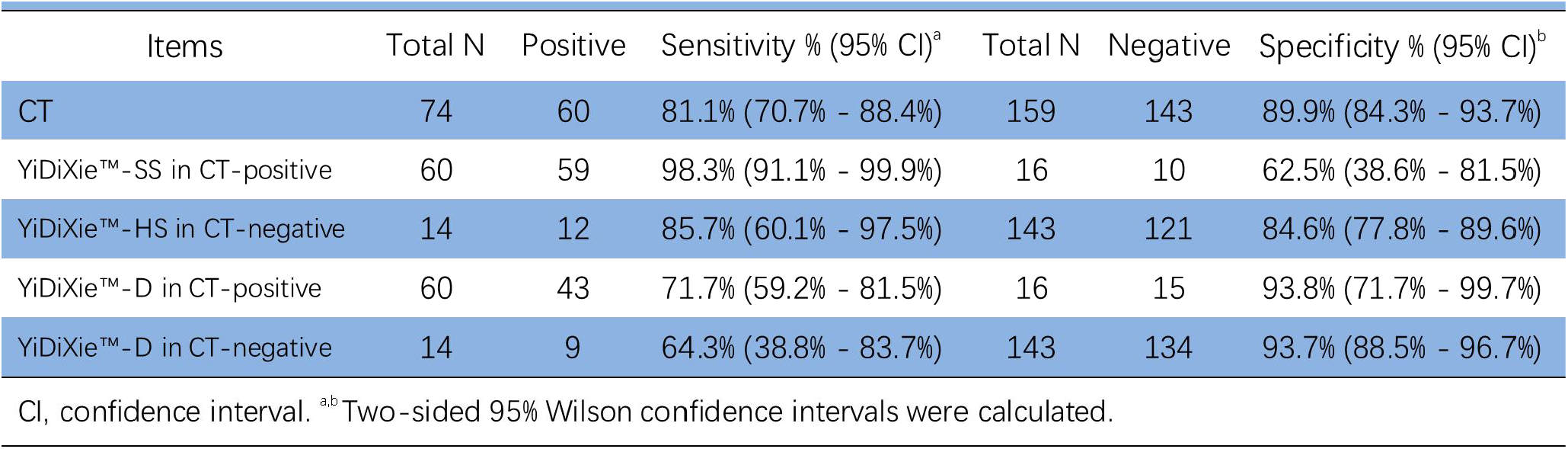
Performance of different Items.

| Items | Total N | Positive | Sensitivity % (95% CI)<sup>a</sup> | Total N | Negative | Specificity % (95% CI)<sup>b</sup> |
| --- | --- | --- | --- | --- | --- | --- |
| CT | 74 | 60 | 81.1% (70.7% - 88.4%) | 159 | 143 | 89.9% (84.3% - 93.7%) |
| YiDiXie™-SS in CT-positive | 60 | 59 | 98.3% (91.1% - 99.9%) | 16 | 10 | 62.5% (38.6% - 81.5%) |
| YiDiXie™-HS in CT-negative | 14 | 12 | 85.7% (60.1% - 97.5%) | 143 | 121 | 84.6% (77.8% - 89.6%) |
| YiDiXie™-D in CT-positive | 60 | 43 | 71.7% (59.2% - 81.5%) | 16 | 15 | 93.8% (71.7% - 99.7%) |
| YiDiXie™-D in CT-negative | 14 | 9 | 64.3% (38.8% - 83.7%) | 143 | 134 | 93.7% (88.5% - 96.7%) |

CI, confidence interval. <sup>a,b</sup>Two-sided 95% Wilson confidence intervals were calculated.

### Diagnostic Performance of YiDiXie™-SS in enhanced MRI-positive patients

To address the challenge of high false-positive rate of brain enhanced MRI, YiDiXie ™ -SS was applied to enhanced MRI-positive patients.

As shown in Table 5, the sensitivity of YiDiXie™ -SS in enhanced MRI-positive patients was 98.3% (95% CI: 91.1% - 99.9%) and its specificity was 62.5% (95% CI: 38.6% - 81.5%).This means that the application of YiDiXie ™ -SS reduces the false-positive rate of brain enhanced MRI by 62.5% (95% CI: 38.6% - 81.5%) with essentially no increase in malignancy leakage.

### Diagnostic Performance of YiDiXie™-HS in enhanced MRI-negative patients

To address the challenge of high false-negative rate of brain enhanced MRI, YiDiXie™ -HS was applied to enhanced MRI-negative patients.

As shown in Table 5, the sensitivity of YiDiXie™ -HS in enhanced MRI-negative patients was 85.7% (95% CI: 60.1% - 97.5%) and its specificity was 84.6% (95% CI: 77.8% - 89.6%).This means that the application of YiDiXie ™ -HS reduces the false-negative rate of brain enhanced MRI by 85.7% (95% CI: 60.1% - 97.5%).

### Diagnostic performance of YiDiXie™-D in enhanced MRI-positive patients

The false-positive consequences of certain brain tumors are significantly more severe than the false-negative consequences, so YiDiXie ™ -D was applied to such patients to reduce their false-positive rates.

As shown in Table 5, YiDiXie ™ -D had a sensitivity of 71.7% (95% CI: 59.2% - 81.5%) and its specificity was 93.8% (95% CI: 71.7% - 99.7%) in patients with positive enhanced MRI.This means that YiDiXie™-SS reduces the false positive rate of enhanced MRI by 93.8% (95% CI: 71.7% - 99.7%).

### Diagnostic Performance of YiDiXie™-D in enhanced MRI-negative patients

The false-positive consequences are significantly more severe than the false-negative consequences in certain enhanced MRI-negative patients, so the more specific YiDiXie ™ -D should be applied to such patients.

As shown in Table 5, YiDiXie ™ -D had a sensitivity of 64.3% (95% CI: 38.8% - 83.7%) and a specificity of 93.7% (95% CI: 88.5% - 96.7%) in patients with negative enhanced MRI. This means that YiDiXie™-D reduces the false-negative rate of enhanced MRI by 64.3% (95% CI: 38.8% - 83.7%) while maintaining high specificity.

## DISCUSSION

### Clinical significance of YiDiXie™-SS in enhanced MRI-positive patients

For patients with positive brain-enhanced MRI, both the sensitivity and specificity of further diagnostic approaches are important. A higher false-negative rate means that more malignant tumors are underdiagnosed, which will lead to a delay in their treatment, progression of the malignant tumor, and possibly even development of advanced stages. Higher false-positive rates mean more benign tumors are misdiagnosed and will likely lead to unnecessary surgery or invasive treatment.

Weighing both the sensitivity and specificity is essentially a trade-off between the “ harm of malignant tumors missed” and the “harm of benign diseases misdiagnosed “. Generally speaking, the misdiagnosis of benign brain tumors as malignant tumors will usually lead to unnecessary surgery or invasive treatment, without affecting the patient’s prognosis, and the cost of treatment is much lower than that of advanced cancer. Thus, the risk of underdiagnosis of malignant tumors in the brain due to false negatives is much greater than the risk of misdiagnosis of benign diseases due to false positives. Furthermore, the positive predictive value was higher in patients with positive brain enhancement MRI. Even if the false-negative rate is comparable to the false-positive rate, the harm is greater. Thus, YiDiXie ™ -SS, which has very high sensitivity but slightly lower specificity, was chosen to reduce the false-positive rate of MRI scans of brain tumors.

As shown in Table 5, YiDiXie ™ -SS had a sensitivity of 98.3%(95% CI: 91.1% - 99.9%) and its specificity was 62.5% (95% CI: 38.6% - 81.5%) in patients with positive enhanced MRI. The above results suggest that YiDiXie ™ -SS reduces the false-positive rate of brain-enhanced MRI by 62.5% (95% CI: 38.6% - 81.5%) while maintaining a sensitivity close to 100%.

Shortly, YiDiXie™-SS substantially reduces the risk of misdiagnosis of benign tumors in patients with false-positive brain-enhanced MRI with essentially no increase in delayed treatment of malignant tumors. Consequently, YiDiXie ™ -SS satisfies the clinical needs well and possesses important clinical significance and a wide range of application prospects.

### Clinical significance of YiDiXie™-HS in enhanced MRI-negative patients

For patients with negative enhancement MRI, further diagnostic methods of both the sensitivity and specificity are important. A higher false-negative rate means that more malignant tumors are missed, which will lead to a delay in their treatment, progression of malignant tumors, and possibly even development of advanced stages. Consequently, the patients will have to bear the adverse consequences of poor prognosis, short survival, poor quality of life, and high cost of treatment. A higher false-positive rate means more benign tumors are misdiagnosed, which will likely lead to unnecessary surgery or invasive treatment.

Weighing both the sensitivity and specificity is essentially a trade-off between the “ danger of malignant tumors being missed” and the “danger of benign tumors being misdiagnosed “. Generally speaking, misdiagnosis of benign brain tumors as malignant tumors will usually lead to unnecessary surgery or invasive treatment, which will not affect the patient’s prognosis, and the cost of treatment is much lower than that of advanced cancers. Thus, a higher false-negative rate results in a greater risk of “underdiagnosis of malignant tumors” in the brain. In addition, the negative predictive value is higher in patients with negative brain enhanced MRI. The higher false-positive rate results in a greater risk of misdiagnosis of benign diseases. Thus, the highly sensitive and specific YiDiXie ™-HS was chosen to reduce the false negative rate of brain enhanced MRI.

As shown in Table 5, YiDiXie ™ -HS had a sensitivity of 85.7% (95% CI: 60.1% - 97.5%) and a specificity of 84.6%(95% CI: 77.8% - 89.6%) in patients with negative enhanced MRI. The above results suggest that the application of YiDiXie ™ -HS reduced the false-negative rate of enhanced MRI by 85.7% (95% CI: 60.1% - 97.5%).

Briefly, YiDiXie ™ -HS significantly reduces the risk of malignant tumor underdiagnosis in patients with false-negative brain-enhanced MRI. Consequently, YiDiXie ™ -HS fulfills the clinical needs well, and is of great clinical significance and broad application prospects.

### Diagnostic performance of YiDiXie™-D

Brain tumors considered malignant are usually treated with surgery or conservative treatments with more side effects. Nevertheless, in some cases, the choice of surgery or invasive treatment needs to be made with extra caution, and therefore further diagnosis is required, e.g., smaller tumors, tumors involving important functional areas of the brain, patients in poor general condition, etc..

Both the sensitivity and specificity of further diagnostic methods are important in patients with brain tumors. Trading off both the sensitivity and specificity is essentially a trade-off between the “ danger of underdiagnosis of malignant tumors “ and the “danger of misdiagnosis of benign tumors”. As smaller tumors have a lower risk of tumor progression and distant metastasis, the risk of underdiagnosis of malignant tumors due to false negatives is much lower than the risk of misdiagnosis of benign tumors due to false positives. In tumors that involve important functional areas of the brain, because surgery or invasive treatment may damage important brain functions, the risk of misdiagnosis of benign tumors due to false positives is much higher than the risk of misdiagnosis of malignant tumors due to false negatives. In patients with poor general conditions, the perioperative risk is much higher than the general condition, and therefore the risk of misdiagnosis of benign tumors due to false positives is much higher than the risk of malignant tumors due to false negatives. Therefore, for these patients, YiDiXie ™ -D, which has a very high specificity but a slightly lower sensitivity, was chosen to reduce the false-positive rate of brain-enhanced MRI or to significantly reduce its false-negative rate while maintaining a high specificity.

As shown in Table 5, the sensitivity of YiDiXie™ -D in patients with positive enhanced MRI was 71.7% (95% CI: 59.2% - 81.5%), and its specificity was 93.8% (95% CI: 71.7% - 99.7%); the sensitivity of YiDiXie™-D in patients with negative enhanced MRI was 64.3% (95% CI: 38.8% - 83.7%). 83.7%) and its specificity was 93.7% (95% CI: 88.5% - 96.7%). These results suggest that YiDiXie ™ -D reduces the false-positive rate of enhanced MRI by 93.8% (95% CI: 71.7% - 99.7%) or reduces the false-negative rate of enhanced MRI by 64.3% (95% CI: 38.8% - 83.7%) while maintaining a high specificity.

In brief, YiDiXie ™-D dramatically reduces the risk of unnecessary surgery or serious perioperative complications in these patients. As such, YiDiXie™ -D well meets the clinical needs and possesses important clinical significance and a wide range of application prospects.

### The YiDiXie™ test are expected to solve 2 challenges in brain tumors

First, YiDiXie ™ -SS can reduce the risk of misdiagnosis of benign brain tumors as malignant tumors. On the one hand, YiDiXie™-SS drastically reduces the risk of misdiagnosis of benign brain tumors with surgery or invasive treatment. Patients with positive brain enhancement MRIs usually undergo surgery or invasive treatment. Because of the high rate of false-positive enhanced MRIs, a large number of patients with benign brain tumors are subjected to unnecessary surgery or invasive treatment. As shown in Table 5, YiDiXie ™ -SS reduces the false-positive rate of enhanced MRI of the brain by 62.5% (95% CI: 38.6% - 81.5%) with essentially no increase in malignant tumor leakage. Therefore, YiDiXie ™ -SS substantially reduces the risk of incorrect surgery or invasive treatment in patients with benign brain tumors without increasing malignant tumor delay in treatment.

On the other hand, YiDiXie™-SS considerably reduces the stress of non-essential work for clinicians. When a brain enhanced MRI is positive, surgery or invasive treatment is usually performed on that patient. Their timely completion is directly dependent on the number of clinicians. In many parts of the world, months or even more than a year’s worth of appointments are available. Inevitably, this delays the treatment of malignant cases among them, thus it is not uncommon for patients with enhanced MRI-positive brain tumors awaiting surgical or invasive treatment to develop malignant progression or even distant metastases. As shown in Table 5, YiDiXie ™ -SS reduces the false-positive rate of brain-enhanced MRI by 62.5% (95% CI: 38.6% - 81.5%) with essentially no increase in malignant tumor leakage. Accordingly, YiDiXie™ -SS significantly relieves clinicians of non-essential workload and promotes timely diagnosis and treatment of malignant tumors or other diseases of the brain that would otherwise be delayed in treatment.

Second, YiDiXie™-HS greatly reduces the risk of underdiagnosis of malignant tumors of the brain. When an enhanced MRI is negative, it usually rules out the possibility of a brain malignancy for the time being. The higher rate of false-negative enhanced MRI results in delayed treatment for a large number of patients with brain malignant tumors. As shown in Table 5, YiDiXie™-HS reduced the false negative rate of enhanced MRI by 85.7% (95% CI: 60.1% - 97.5%). Therefore, YiDiXie ™ -HS substantially reduces the probability of false-negative enhanced MRI miss-diagnosis of malignant tumors and facilitates timely diagnosis and treatment of patients with malignant tumors of the brain who would otherwise be delayed in treatment.

Third, YiDiXie ™ -D is expected to meet the challenges of “high false positive rate” and “high false negative rate” in some specific patients. Brain tumors that are considered malignant are usually treated with surgery or conservative treatments with significant side effects. However, in cases such as small tumors, tumors accumulating important functional areas of the brain, and patients with poor general conditions, surgery or invasive treatment may lead to unnecessary surgical trauma or serious perioperative complications. Therefore these patients require extra caution before surgery or invasive treatment. As shown in Table 5, YiDiXie ™-SS reduced the false-positive rate of enhanced MRI by 93.8% (95% CI: 71.7% - 99.7%) or the false-negative rate of enhanced MRI by 64.3% (95% CI: 38.8% - 83.7%) while maintaining a high specificity. As a result, YiDiXie ™ -D dramatically reduces the risk of incorrect surgery, serious perioperative complications or severe treatment side effects in these patients.

Final, The YiDiXie™ test enables “just-in-time” diagnosis of brain tumors. On the one hand, the YiDiXie ™ test only requires microscopic amounts of blood, enabling patients to complete the diagnostic process non-invasively without having to leave their homes. A single the YiDiXie ™ test requires only 20 microliters of serum, which is approximately the same amount as 1 drop of whole blood^8^. Given the pre-test sample quality assessment experiments and 2-3 repeat experiments, 0.2 ml of whole blood is adequate to perform the YiDiXie ™ test^8^. The average patient can use a finger blood collection needle to collect 0.2 ml of finger blood at home, instead of requiring venous blood collection by medical personnel, making the diagnostic process non-invasive without having to leave the patient’s home^8^.

On the other hand, the YiDiXie ™ test has a nearly unlimited diagnostic capacity. Figure 1 shows the basic flow chart of the YiDiXie ™ test, which shows that the YiDiXie ™ test not only requires no doctors or medical equipment, but also requires no medical personnel to collect blood^8^. Therefore, the YiDiXie ™ test is completely independent of the number of medical personnel and medical facilities, and its testing capacity is nearly unlimited^8^. This allows the YiDiXie™ test to “ just-in-time” diagnose brain tumors without the patient having to wait anxiously for an appointment.

**Figure 1.**
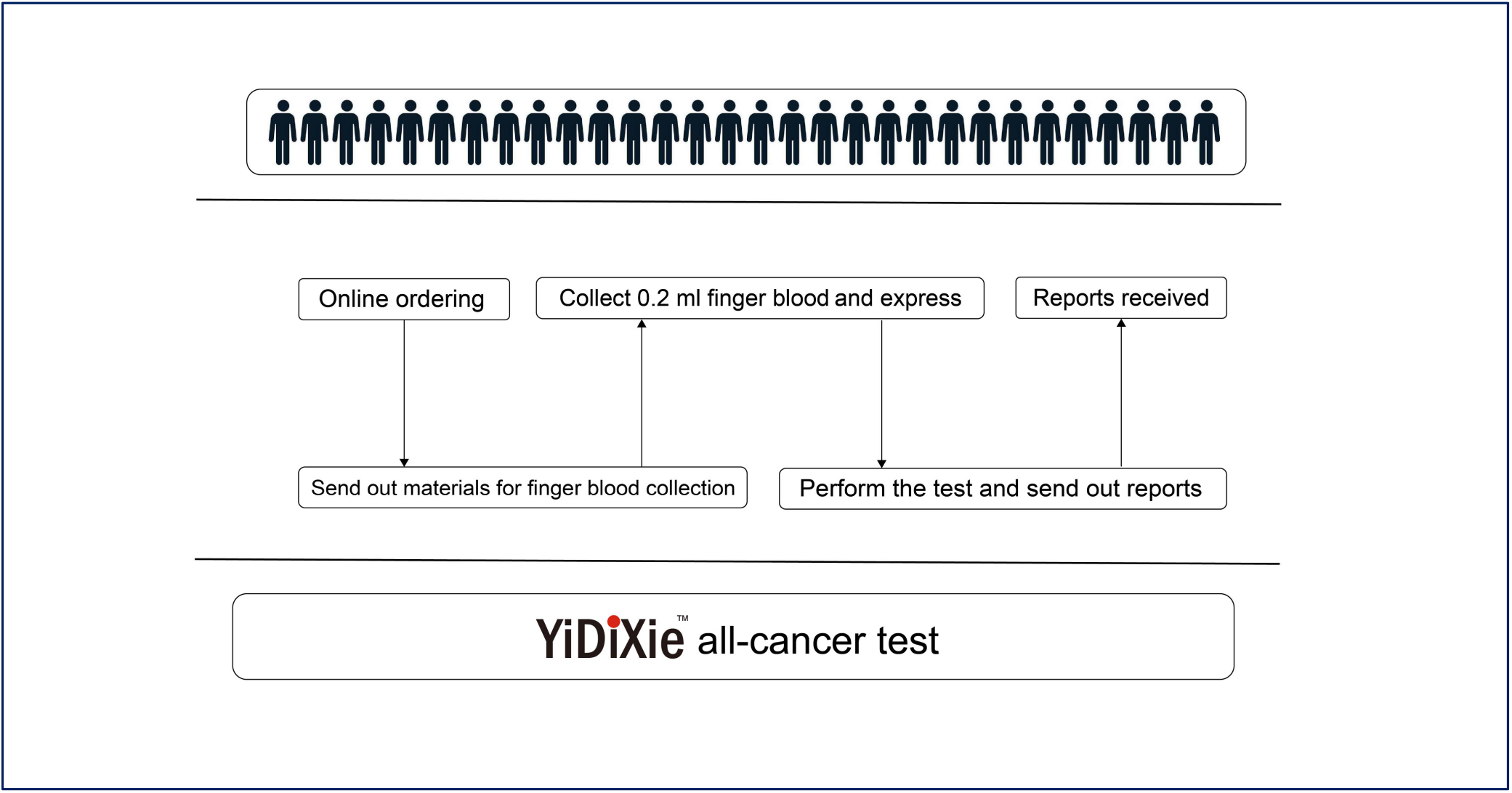
Basic flowchart of the YiDiXie™ test.

In short, the YiDiXie ™ test has an important diagnostic value in brain tumors, and it is expected to solve the problems of “high false-negative rate of enhanced MRI” and “high false-positive rate of enhanced MRI” for brain tumors.

### Limitations of the study

First, this study had a small number of cases and future clinical studies with larger sample sizes are needed for further evaluation.

Second, the present study was an inpatient malignant tumor case-benign tumor control study, and future cohort studies in the natural population of brain tumors are needed for further evaluation.

Final, the current study was a single-center study, which may have led to some degree of bias in the results of this study. Multicenter studies are needed to further evaluate this in the future.

## CONCLUSION

YiDiXie™-SS has extremely high sensitivity and high specificity in brain tumors.YiDiXie ™ -HS has high sensitivity and high specificity in brain tumors.YiDiXie ™ -D has high sensitivity and extremely high specificity in brain tumors.YiDiXie™ -SS significantly reduces the false-positive rate of brain-enhanced MRIs with essentially no increase in delayed treatment of malignant tumors. The YiDiXie ™ -HS significantly reduces the false-negative rate of brain-enhanced MRI. the YiDiXie ™ -D can significantly reduce the false-positive rate of brain-enhanced MRI or significantly reduce the false-negative rate of brain-enhanced MRI while maintaining a high level of specificity. The YiDiXie ™ test has significant diagnostic value in brain tumors, and is expected to solve the problems of “ high false-positive rate “ and “high false-negative rate” of brain-enhanced MRI.

## Data Availability

All data produced in the present study are contained in the manuscript.

## FUNDING

This study was supported by Shenzhen High-level Hospital Construction Fund, Clinical Research Project of Peking University Shenzhen Hospital (LCYJ2020002, LCYJ2020015, LCYJ2020020, LCYJ2017001).

## Notes

### Competing Interest Statement

The authors have declared no competing interest.

### Clinical Trial

ChiCTR2200066840

### Funding Statement

This work was supported by Shenzhen High-level Hospital Construction Fund, Clinical Research Project of Peking University Shenzhen Hospital (LCYJ2020002, LCYJ2020015, LCYJ2020020, LCYJ2017001).

### Author Declarations

Ethics committee of Peking University Shenzhen Hospital gave ethical approval for this work.

### Summary of Updates

The results were updated. Tabel 2-8 were reviesed.

